# COVID-19 Prevention Beliefs and Practices in College Students

**DOI:** 10.1101/2021.01.29.21250794

**Authors:** Caitlyn Cassimatis, Liga Kreitner, Abdoulie Njai, Emily Leary, Aaron Gray

**Affiliations:** School of Medicine, University of Missouri, Columbia, MO, USA; Department of Orthopaedic Surgery, University of Missouri, Columbia, MO, USA; Department of Clinical Family and Community Medicine, University of Missouri, Columbia, MO, USA; Thompson Laboratory for Regenerative Orthopaedics, University of Missouri, Columbia, MO, USA

## Abstract

**Background:** As college students in the United States return to university campuses, it is important to understand their beliefs and practices on coronavirus disease 2019 (COVID-19) prevention.

**Purpose:** To assess beliefs and practices regarding COVID-19 prevention among college students in the United States

**Methods:** An online, self-administered survey was developed that collected information on COVID-19 preventative practice and beliefs. Survey responses were collected between July 13, 2020 and July 31, 2020.

**Results:** A total of 4,834 college students participated in the survey with a response rate of 22.9%. Compared to males, more female college students practiced COVID-19 preventative measures, including *always* wearing masks or face coverings in public (52% vs. 44%, p<0.001) and *always* or *often* observing social distancing (70% vs. 63%, p<0.001). In contrast to students from larger population areas, fewer college students from rural areas reported practicing prevention measures, such as *always* wearing a mask (24% rural v. 45% towns vs. 55% cities, p<0.001) and *always* social distancing (20% rural vs. 21% towns vs. 29% cities, p<0.001). Additionally, more students from rural areas have become *much less* worried about personally contracting COVID-19 over the last 3 months when compared to students from towns and cities (21% vs. 16% vs. 11%, p<0.001). Fewer white college students compared to other racial groups thought it was *very important* to wear masks (55% white vs. 76% Black vs. 82% Asian vs. 63% American Indian or Alaskan native (AIAN) & Native Hawaiian or Other Pacific Islander (NHOPI), p<0.001) and *very important* to practice social distancing (29% white vs. 50% Black vs. 53% Asian vs. 36% AIAN/NHOPI, p <0.001). Compared to Non-Hispanic students, more Hispanic students thought it was *very important* to practice preventative measures, including wearing a mask (71% vs. 58%, p<0.001), social distancing (37% vs. 32%, p=0.017), and good hand hygiene (77% vs. 67%, p=0.013).

**Conclusion:** COVID-19 prevention beliefs and practices differ between sexes, the size of town one lives, race, and ethnicity. In general, female students followed Center for Disease Control and Prevention (CDC) COVID-19 prevention guidelines more closely than male students. Students who reside in areas of larger populations have more strict COVID-19 prevention beliefs and practices than students from areas with smaller populations. Asian and Black/African American students adhered closer to CDC COVID-19 prevention guidelines and had stronger beliefs for infection prevention measures than white or AIAN/NHOPI students. Hispanic/Latino students were more stringent in COVID-19 prevention beliefs and practices than non-Hispanic/Latino students.

## INTRODUCTION

In late 2019, a novel coronavirus that generally presents as an acute viral respiratory tract infection was first identified in Wuhan, China.^1^ Coronavirus disease 2019 (COVID-19) quickly spread to other countries around the world. On March 11, 2020, the World Health Organization (WHO) characterized coronavirus disease 2019 (COVID-19) as a pandemic.^2^

There is currently no Food and Drug Administration approved vaccine for COVID-19, so it is important that the public understand and implement preventative measures. To educate the U.S. public on how to prevent the spread of COVID-19, the Centers for Disease Control and Prevention (CDC) developed guidelines entitled “How to Protect Yourself and Others”, which outline preventative practices such as washing hands often, avoiding close contact with others (6 feet of distance), and wearing a cloth face cover when around others.^3^ A study involving Pakistani university students conducted in February and March, 2020 found that students had good knowledge and attitudes about COVID-19, but poor preventative practices.^4^

As colleges and universities resume classes, it is important to understand students’ beliefs about COVID-19 prevention and how they practice preventative measures. This survey was designed to assess beliefs and practices on COVID-19 prevention among college students in the United States.

## METHODS

All procedures performed in this study were administered with institutional review board approval. All undergraduates enrolled at a single large university in the Midwest for the Fall 2020 semester were invited to participate. Undergraduate students were emailed directly by the researchers to request participation in the study. There was no in-person subject interaction. Before beginning the survey, participants were given study information and an electronic consent form. The email contained a description of the study and a link to the online, anonymous survey. In order to participate, students were required to confirm that they met the eligibility criteria and agreed to participate. Eligible participants included undergraduate students, ages 18 years and older, enrolled for the Fall 2020 semester at the participating university. Survey responses were collected between July 13, 2020 and July 31, 2020.

### COVID-19 Prevention Beliefs and Practices

To assess COVID-19 prevention beliefs and practices, participants answered a series of survey questions. These questions were developed using the CDC COVID-19 guidelines on “How to Protect Yourself & Others”.^3^ To assess COVID-19 prevention *beliefs*, participants were asked whether they thought it was important to wear a mask or face covering in public places, stay 6 feet apart (i.e., social distancing) from others, and frequently wash their hands or use hand sanitizer. Items to assess participants’ COVID-19 prevention *practices* included how often they wore a mask or face covering when going to the grocery store since March 2020 and how often they tried to stay 6 feet apart (i.e., social distancing) from people who were not family or roommates. Participants also answered questions to assess how worried they were about personally catching COVID-19, about a family member catching COVID-19, and if they had become more or less worried about catching COVID-19 over the past 3 months.

### Demographics

Participants completed a brief set of demographic questions. These questions included information on race, ethnicity, sex, gender, state lived in most since mid-March 2020, population of the town or city lived in most since mid-March 2020, and if they are currently living in the same town as their university. We collected sex and gender information using two-step gender identity questions previously described by Bauer et al.^5^ Mid-March 2020 was chosen as the reference date as that is when the WHO officially classified COVID-19 as a pandemic.^2^ In addition, race was recorded as “any applicable” from the list of six, which was aggregated for analysis. Race responses were collapsed as outlined in Supplemental Table 1.

### Statistical Analysis

Survey data were summarized and aggregated. Bivariate relationships were determined using the two-sided Chi-square test for association and Fisher’s Exact test, where appropriate. Stratified analyses were considered, but there were small samples sizes in cohorts that prevented statistical assumptions from being met for stratified analysis. Significant p-values were set at p<0.05.

## RESULTS

An email with the project description and survey link was sent to 21,128 undergraduate students. Of these, 4,834 students consented to participate and met inclusion criteria for a response rate of 22.9%.

### Differences in Sex and Gender

In general, female students followed CDC COVID-19 prevention guidelines more closely than male students (Table 2.) Since March 2020, 52% of females vs 44% of males (p<0.001) have *always* worn a mask or face covering when going to a grocery store, 31% of males vs 18% of females (p<0.001) were *not worried* about personally catching COVID-19, and 11% of females vs. 18 percent of males (p<0.001) had become *much less worried* about catching COVID-19 over the past 3 months. 70% of females vs 63% of males (p<0.001) *always* or *often* tried to stay 6 feet apart (i.e, social distancing) from people who are not family or roommates when out in public. On the topic of hand sanitizing, 74% of females vs 56% of males (p<0.001) think that it is *very important* to frequently wash their hands with soap and water for at least 20 seconds or use hand sanitizer. Self-reported current gender identity was asked of participants, but too few responses other than male or female were collected (1.1%), so analyses focused on sex assigned at birth.

**Table 1.**
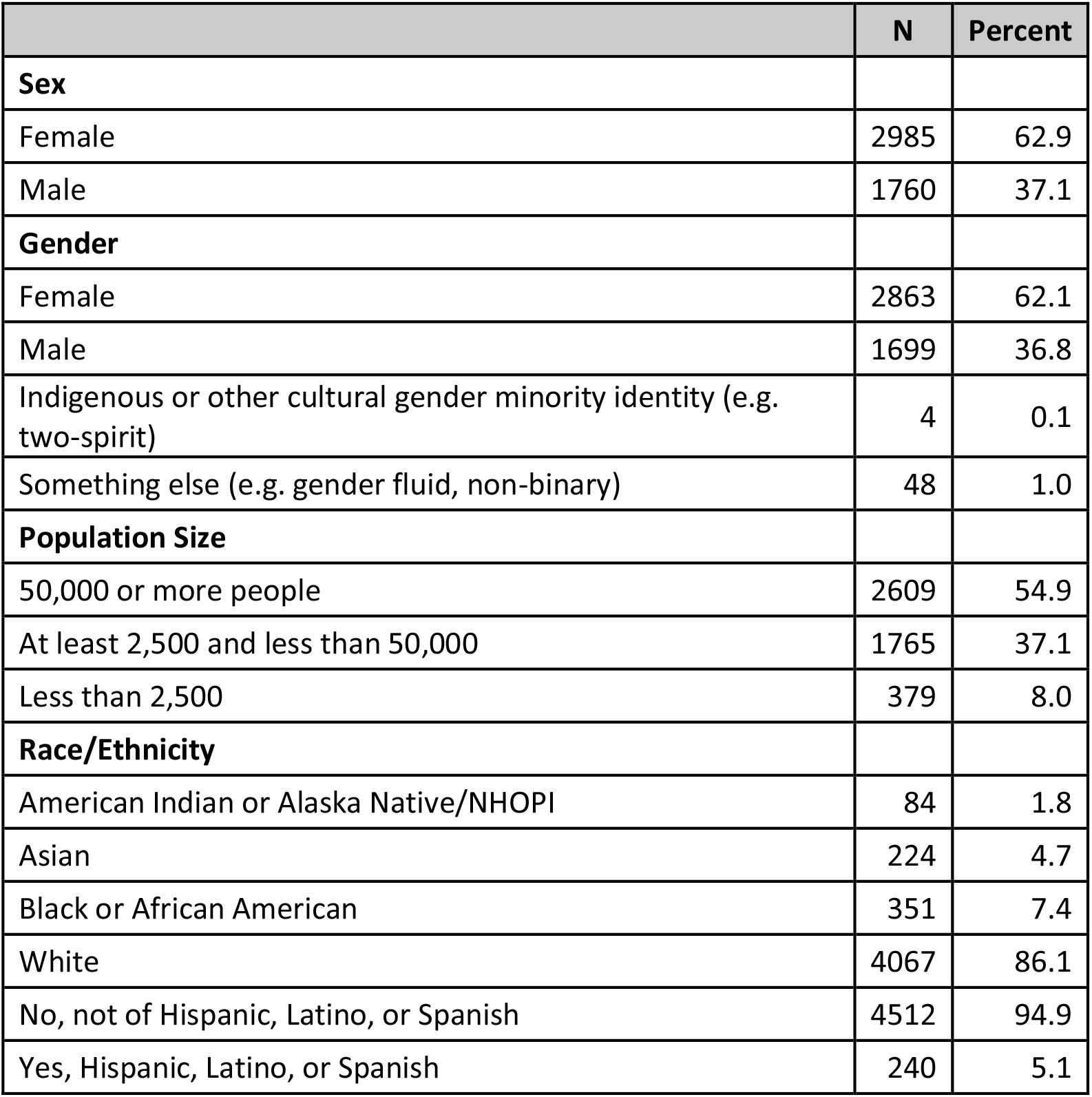
Summary statistics for variables of interest (grouping variables)

|  | N | Percent |
| --- | --- | --- |
| **Sex** |  |  |
| Female | 2985 | 62.9 |
| Male | 1760 | 37.1 |
| **Gender** |  |  |
| Female | 2863 | 62.1 |
| Male | 1699 | 36.8 |
| Indigenous or other cultural gender minority identity (e.g. two-spirit) | 4 | 0.1 |
| Something else (e.g. gender fluid, non-binary) | 48 | 1.0 |
| **Population Size** |  |  |
| 50,000 or more people | 2609 | 54.9 |
| At least 2,500 and less than 50,000 | 1765 | 37.1 |
| Less than 2,500 | 379 | 8.0 |
| **Race/Ethnicity** |  |  |
| American Indian or Alaska Native/NHOPI | 84 | 1.8 |
| Asian | 224 | 4.7 |
| Black or African American | 351 | 7.4 |
| White | 4067 | 86.1 |
| No, not of Hispanic, Latino, or Spanish | 4512 | 94.9 |
| Yes, Hispanic, Latino, or Spanish | 240 | 5.1 |

**Table 2.**
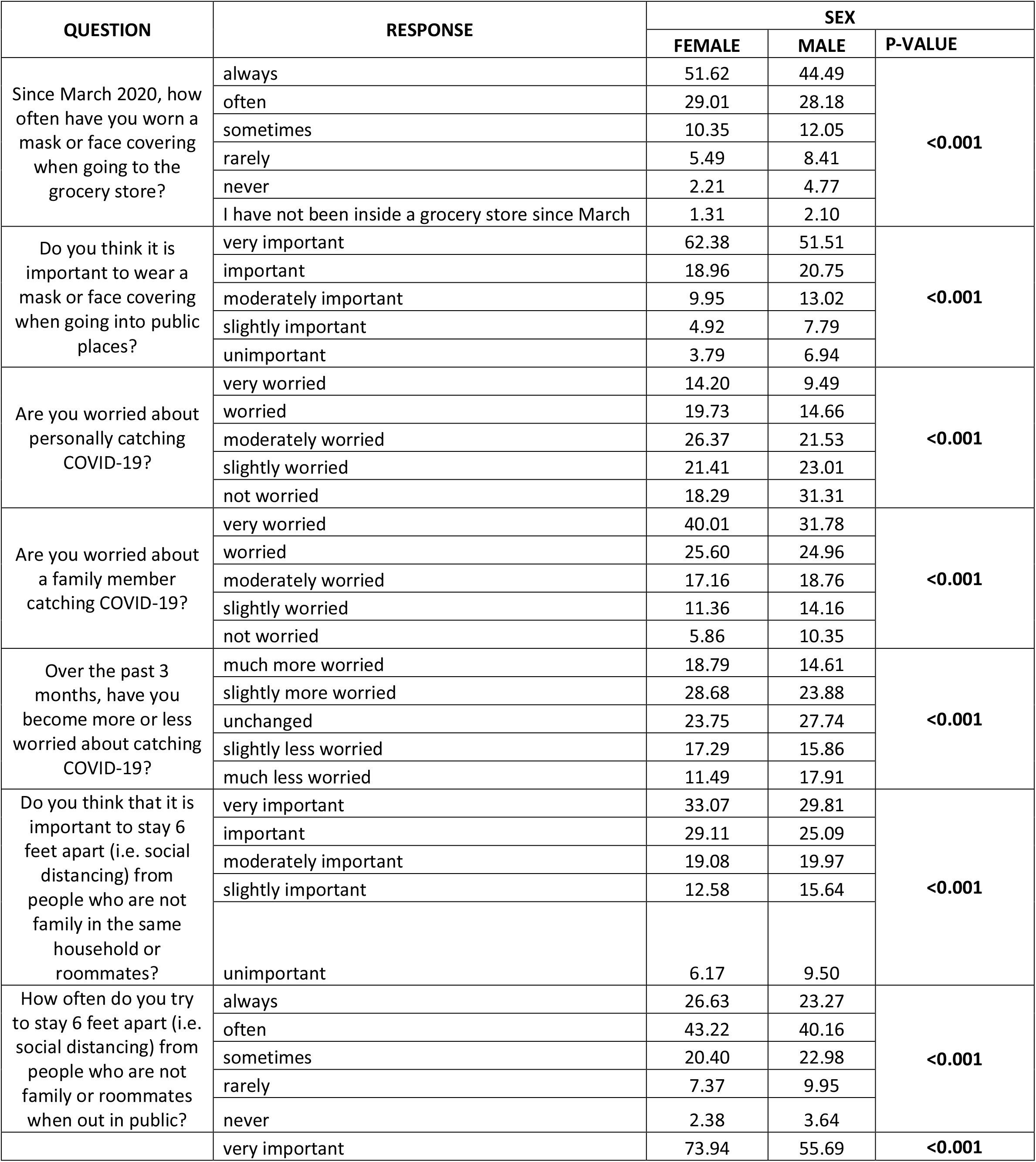
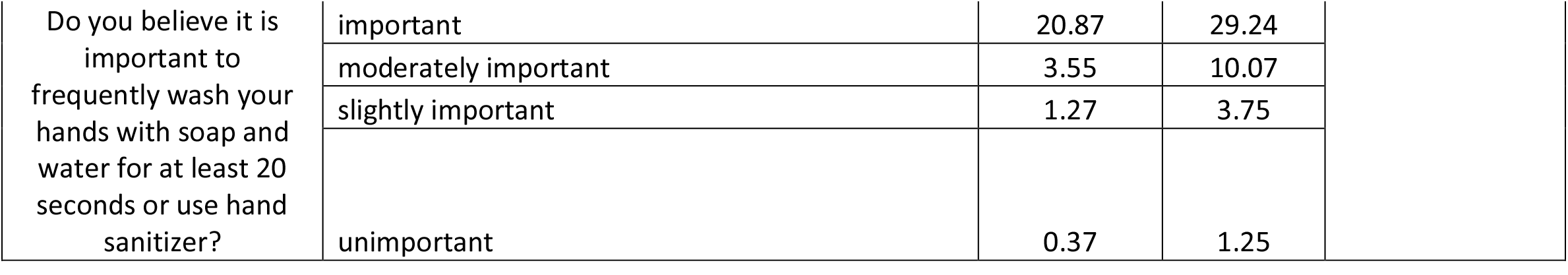
COVID-19 prevention beliefs and practices by sex assigned at birth.

| QUESTION | RESPONSE | SEX |  |  |
| --- | --- | --- | --- | --- |
|  |  | FEMALE | MALE | P-VALUE |
| Since March 2020, how often have you worn a mask or face covering when going to the grocery store? | always | 51.62 | 44.49 | <0.001 |
|  | often | 29.01 | 28.18 |  |
|  | sometimes | 10.35 | 12.05 |  |
|  | rarely | 5.49 | 8.41 |  |
|  | never | 2.21 | 4.77 |  |
|  | I have not been inside a grocery store since March | 1.31 | 2.10 |  |
| Do you think it is important to wear a mask or face covering when going into public places? | very important | 62.38 | 51.51 | <0.001 |
|  | important | 18.96 | 20.75 |  |
|  | moderately important | 9.95 | 13.02 |  |
|  | slightly important | 4.92 | 7.79 |  |
|  | unimportant | 3.79 | 6.94 |  |
| Are you worried about personally catching COVID-19? | very worried | 14.20 | 9.49 | <0.001 |
|  | worried | 19.73 | 14.66 |  |
|  | moderately worried | 26.37 | 21.53 |  |
|  | slightly worried | 21.41 | 23.01 |  |
|  | not worried | 18.29 | 31.31 |  |
| Are you worried about a family member catching COVID-19? | very worried | 40.01 | 31.78 | <0.001 |
|  | worried | 25.60 | 24.96 |  |
|  | moderately worried | 17.16 | 18.76 |  |
|  | slightly worried | 11.36 | 14.16 |  |
|  | not worried | 5.86 | 10.35 |  |
| Over the past 3 months, have you become more or less worried about catching COVID-19? | much more worried | 18.79 | 14.61 | <0.001 |
|  | slightly more worried | 28.68 | 23.88 |  |
|  | unchanged | 23.75 | 27.74 |  |
|  | slightly less worried | 17.29 | 15.86 |  |
|  | much less worried | 11.49 | 17.91 |  |
| Do you think that it is important to stay 6 feet apart (i.e. social distancing) from people who are not family in the same household or roommates? | very important | 33.07 | 29.81 | <0.001 |
|  | important | 29.11 | 25.09 |  |
|  | moderately important | 19.08 | 19.97 |  |
|  | slightly important | 12.58 | 15.64 |  |
|  | unimportant | 6.17 | 9.50 |  |
| How often do you try to stay 6 feet apart (i.e. social distancing) from people who are not family or roommates when out in public? | always | 26.63 | 23.27 | <0.001 |
|  | often | 43.22 | 40.16 |  |
|  | sometimes | 20.40 | 22.98 |  |
|  | rarely | 7.37 | 9.95 |  |
|  | never | 2.38 | 3.64 |  |
|  | very important | 73.94 | 55.69 | <0.001 |

|  |  |  |  |
| --- | --- | --- | --- |
| Do you believe it is important to frequently wash your hands with soap and water for at least 20 seconds or use hand sanitizer? | important | 20.87 | 29.24 |
|  | moderately important | 3.55 | 10.07 |
|  | slightly important | 1.27 | 3.75 |
|  | unimportant | 0.37 | 1.25 |

### Differences in Size of Town

Overall, students who reside in areas of larger populations have more-strict COVID-19 prevention beliefs and practices than students from areas with smaller populations. (Table 3) There appears to be a decreasing pattern of concern of COVID-19 in areas as population size decreases. Participants were asked to identify the population size of the town or city they had mostly resided since mid-March 2020. These are classified as “cities”, which are defined to include populations of greater than 50,000; “towns”, which are defined to include populations between 2,500 and 50,000; and “rural areas”, which are defined as populations less than 2,500.

**Table 3.** COVID-19 prevention beliefs and practices by differences in size of town of residence.

| QUESTION | RESPONSE | SIZE OF CITY |  |  |  |
| --- | --- | --- | --- | --- | --- |
|  |  | CITY | TOWN | RURAL | P-VALUE |
| Since March 2020, how often have you worn a mask or face covering when going to the grocery store? | always | 24.34 | 55.12 | 45.10 | <0.001 |
|  | often | 28.04 | 28.90 | 28.50 |  |
|  | sometimes | 20.37 | 8.74 | 12.46 |  |
|  | rarely | 12.96 | 4.41 | 8.39 |  |
|  | never | 12.70 | 1.23 | 3.97 |  |
|  | I have not been inside a grocery store since March | 1.59 | 1.61 | 1.59 |  |
| Do you think it is important to wear a mask or face covering when going into public places? | very important | 38.36 | 65.95 | 51.44 | <0.001 |
|  | important | 20.63 | 17.87 | 21.93 |  |
|  | moderately important | 17.72 | 8.59 | 13.37 |  |
|  | slightly important | 13.23 | 4.14 | 7.20 |  |
|  | unimportant | 10.05 | 3.45 | 6.06 |  |
| Are you worried about personally catching COVID-19? | very worried | 6.08 | 15.29 | 9.58 | <0.001 |
|  | worried | 13.23 | 19.05 | 17.17 |  |
|  | moderately worried | 22.22 | 25.87 | 23.00 |  |
|  | slightly worried | 26.46 | 19.85 | 24.31 |  |
|  | not worried | 32.01 | 19.93 | 25.95 |  |
| Are you worried about a family member catching COVID-19? | very worried | 28.31 | 41.60 | 31.97 | <0.001 |
|  | worried | 24.87 | 24.96 | 25.96 |  |
|  | moderately worried | 22.49 | 15.64 | 19.95 |  |
|  | slightly worried | 15.61 | 11.20 | 13.44 |  |
|  | not worried | 8.73 | 6.60 | 8.67 |  |
| Over the past 3 months, have you become more or less worried about catching COVID-19? | much more worried | 8.99 | 21.08 | 13.27 | <0.001 |
|  | slightly more worried | 23.02 | 28.59 | 25.40 |  |
|  | unchanged | 30.69 | 23.92 | 25.96 |  |
|  | slightly less worried | 16.67 | 15.02 | 19.22 |  |
|  | much less worried | 20.63 | 11.38 | 16.16 |  |
| Do you think that it is important to stay 6 feet apart (i.e. social distancing) from people who are not family in the same household or roommates? | very important | 21.22 | 37.44 | 25.98 | <0.001 |
|  | important | 25.20 | 27.08 | 28.87 |  |
|  | moderately important | 22.81 | 18.14 | 20.70 |  |
|  | slightly important | 19.36 | 11.32 | 15.94 |  |
|  | unimportant | 11.41 | 6.02 | 8.51 |  |
| How often do you try to stay 6 feet apart (i.e. social distancing) from people who are not family or roommates when out in public? | always | 19.84 | 29.15 | 21.36 | <0.001 |
|  | often | 38.36 | 42.81 | 41.64 |  |
|  | sometimes | 24.34 | 19.49 | 23.34 |  |
|  | rarely | 13.49 | 6.41 | 10.03 |  |
|  | never | 3.97 | 2.15 | 3.63 |  |
| Do you believe it is important to frequently wash your hands with soap and water for at least 20 seconds or use hand sanitizer? | very important | 62.43 | 69.44 | 64.74 | **0.006** |
|  | important | 25.40 | 22.58 | 25.85 |  |
|  | moderately important | 7.41 | 5.64 | 6.12 |  |
|  | slightly important | 3.70 | 1.88 | 2.32 |  |
|  | unimportant | 1.06 | 0.46 | 0.96 |  |

**Table 4.**
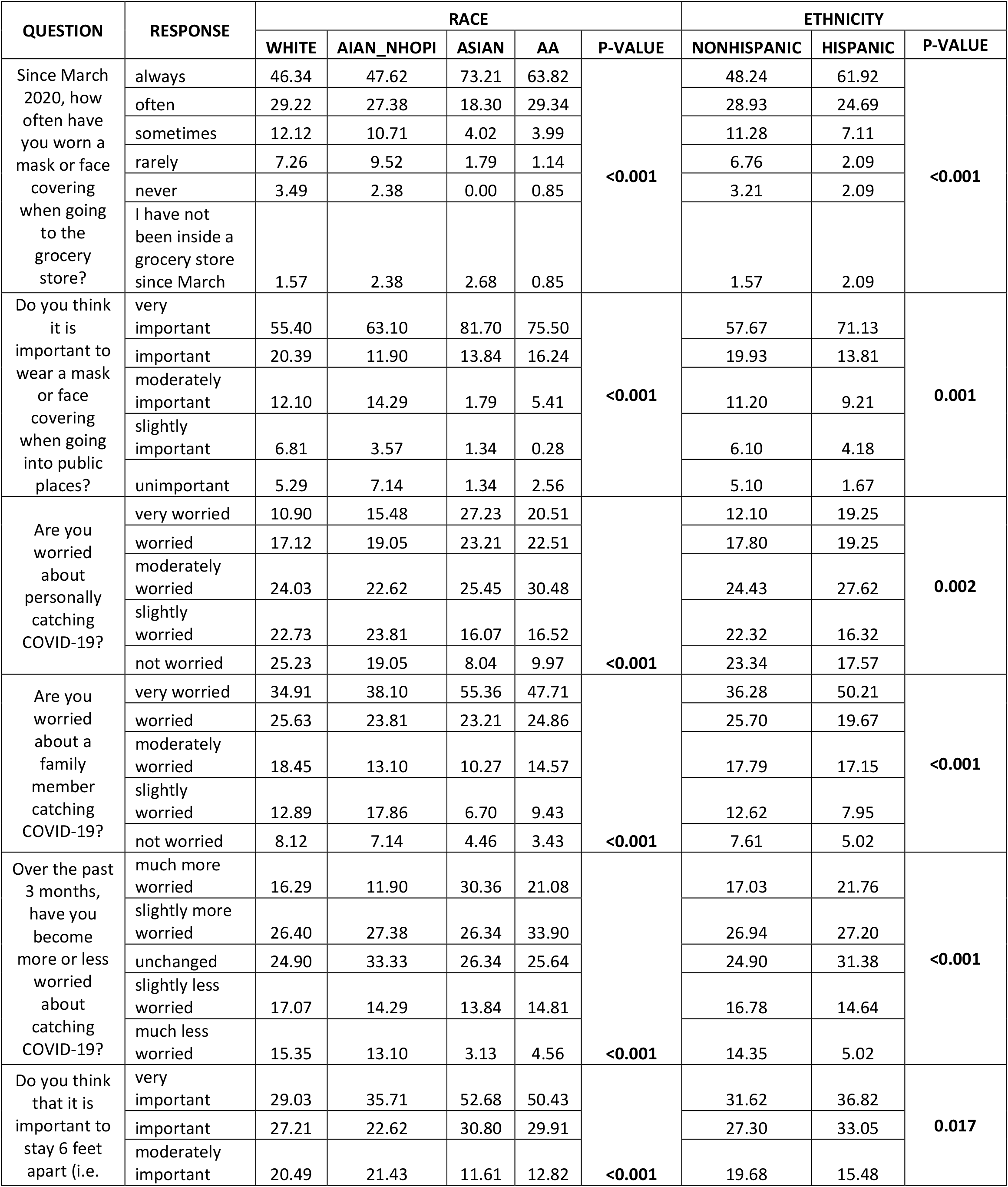
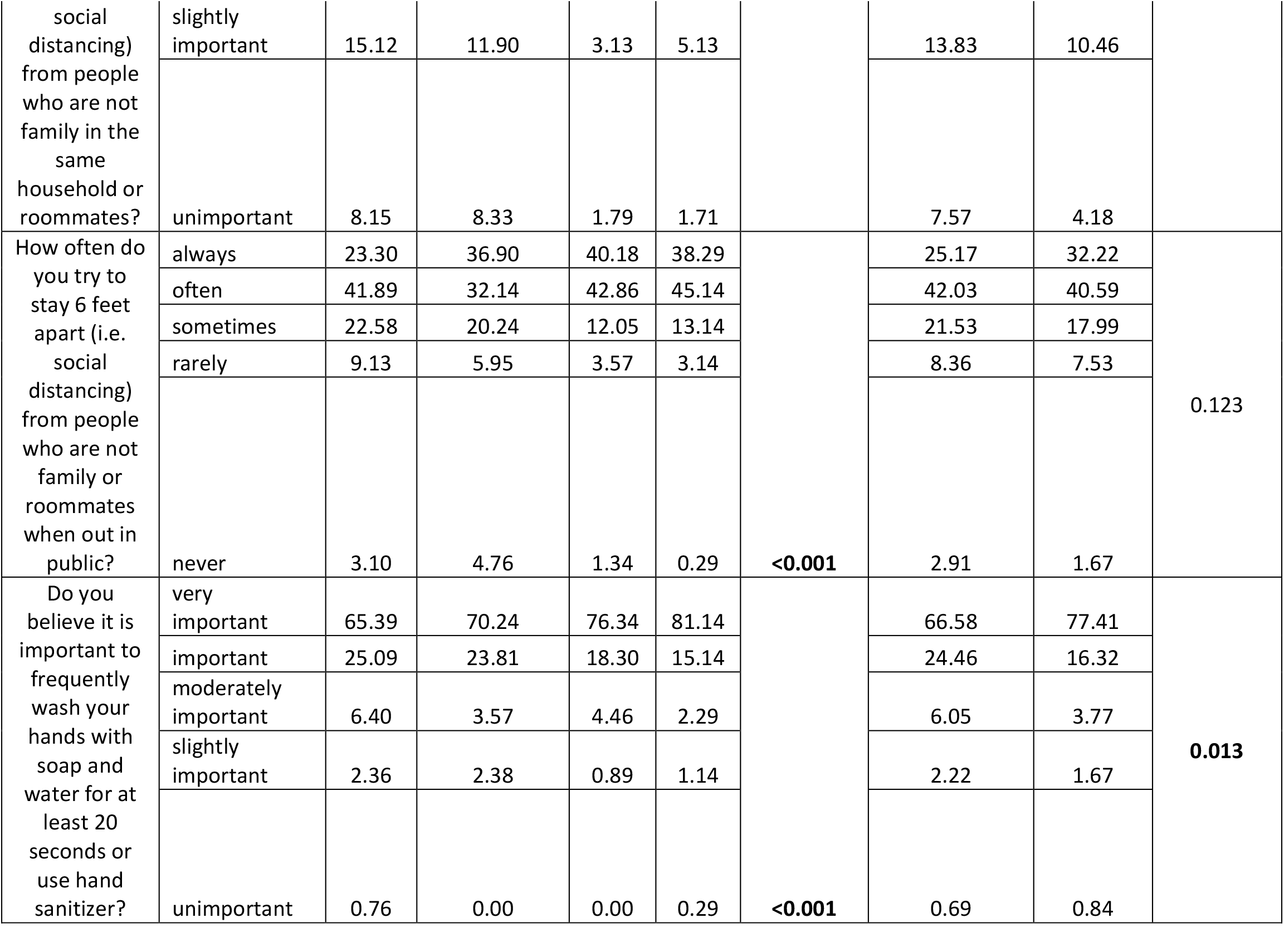
COVID-19 prevention beliefs and practices by race and ethnicity.

City residents (66%) vs 51% in towns vs 38% in rural areas (p<0.001) think it is very important to wear a mask or face covering when going into public places. City residents (42%) vs 32% in towns vs 28% in rural areas (p<0.001) are very worried about a family member catching COVID-19. City residents (21%) vs 13% in towns vs 9% in rural areas (p<0.001) are much more worried now about catching COVID-19 compared to 3 months prior. City residents (37%) vs 26% in towns vs 21% in rural areas (p<0.001) think it is very important to stay 6 feet apart (i.e., social distancing) from people who are not family in the same household or roommates. City residents (69%) vs 65% in towns vs 62% in rural areas (p<0.001) believe it is very important to frequently wash hands with soap and water for at least 20 seconds or use hand sanitizer.

### Differences in Race and Ethnicity

Asian and Black/African American students adhered closer to CDC COVID-19 prevention guidelines and had stronger beliefs for infection prevention measures than white or American Indian or Alaska Native/Native Hawaiian or Other Pacific Islander (AIAN/NHOPI) students. Hispanic/Latino students were more stringent in COVID-19 prevention beliefs and practices than non-Hispanic/Latino students. In general, white, non-Hispanic/Latino students follow CDC COVID-19 prevention guidelines less than other race and ethnicity groups.

When exploring differences by racial groups, 82% of Asian vs 76% of Black vs 63% of AIAN/NHOPI vs 55% of white students (p<0.001) believe it is *very important* to wear a mask or face covering when going into public places, and 25% of white vs 19% of AIAN/NHOPI vs 10% of Black vs 8% of Asian students (p<0.001) are *not worried* about personally catching COVID-19. Students were more worried about their family members becoming sick from COVID-19 than themselves. Of these groups, 55% of Asian vs 48% of Black vs 38% of AIAN/NHOPI vs 35% of white students (p<0.001) are *very worried* about a family member catching COVID-19, and 53% of Asian vs 50% of Black vs 36% of AIAN/NHOPI vs 29% of white students (p<0.001) think it is *very important* to stay 6 feet apart (i.e., social distancing) from people who are not family in the same household or roommates.

When examining COVID-19 prevention practice and belief differences by ethnicity, 71% Hispanic/Latino vs 58% Non-Hispanic/Latino students (p=0.001) believe it is *very important* to wear a mask or face covering when going into public places, while 50% Hispanic/Latino vs 36% Non-Hispanic/Latino students (p<0.001) are *very worried* about a family member catching COVID-19. In this group, 37% Hispanic/Latino vs 32% Non-Hispanic/Latino students (p=0.017) think it is *very important* to stay 6 feet apart (i.e., social distancing) from people who are not family in the same household or roommates, and 77% Hispanic/Latino vs 67% Non-Hispanic/Latino students (p=0.013) think it is *very important* to frequently wash hands with soap and water for at least 20 seconds or use hand sanitizer.

## DISCUSSION

When college students started to return to campuses across the country, many universities experienced cases of COVID-19, leading to quarantines and some transitions to remote learning. While much is still unknown regarding this virus, mitigation strategies like avoiding large gatherings, encouraging mask use, and social distancing have been found to help fight the spread of COVID-19.^2^ As college students around the nation are being encouraged to take personal responsibility for their health, it is important to understand their perspectives about COVID-19 prevention and how closely they followed recommended preventative measures. Overall, we found that COVID-19 prevention beliefs and practices differ significantly between sexes, the size of town one lives in, race, and ethnicity.

### Differences in Sex and Gender

Male students reported less strict prevention beliefs and usage of prevention practices than women. Sex differences exist in mask wearing behavior and some men may have been reluctant to wear them due to viewing masks as “shameful, not cool, and a sign of weakness.”^6^ Female college students reported wearing a mask or face covering in public places at greater rates than males. These results also showed that male students were less concerned than female students about personally contracting COVID-19 or having a family member become ill from COVID-19. Societal gender norms likely play a role in the gender-based differences observed.^7^ Men are conditioned at a young age to be tough, stoic, engage in risky behaviors, and show no weakness.^7^ These characteristics have been linked to men driving at higher speeds, binge drink at higher rates, and being less likely to wear seatbelts.^8^

### Differences in Size of Town

Fewer college students living in rural areas reported concern about COVID-19 infection compared to students living in larger cities and towns. Additionally, students from rural areas reported a lower adherence to CDC preventative recommendations compared to students from towns and cities with higher populations. These findings are similar to a previous study by Czeisler et al. that found that a higher percentage of individuals from urban areas reported use of cloth face coverings compared to individuals from rural areas in the United States.^9^ Differences in preventative practices between individuals in rural and urban communities may be influenced by a number of factors including local policies and transmission rates as well as health literacy.^10^

State and city policies regarding COVID-19 prevention and mitigation strategies, such as stay-at-home orders, nonessential business closures, and public mask mandates, vary widely between local governments in the United States. With a few exceptions, rural areas were less affected by COVID-19 early in the pandemic compared to large metropolitan areas.^11,12^ As a result, rural areas may have been less likely to implement government-mandated prevention strategies to reduce the spread of COVID-19. Lack of these local preventative and mitigation strategies as well as lower prevalence of COVID-19 in rural areas may decrease residents concern for the virus and contribute to a lower likelihood of following CDC recommendations on COVID-19 prevention.

In addition to local policy, decreased socioeconomic resources in rural areas can contribute to lower health literacy in these locations.^13^ COVID-19 prevention strategies require that individuals possess sufficient knowledge on how to reduce their risk of infection through measures such as good hand hygiene, social distancing, and wearing a mask. Without adequate awareness of proper prevention strategies and their importance in reducing viral transmission, residents in rural communities may be less likely to adhere to CDC guidance.

Prevention of COVID-19 transmission in rural areas is especially important as disparities in health conditions and limited healthcare infrastructure increases the consequences of COVID-19 infection.^13^ Rural populations have a higher prevalence of diseases that increase their risk of severe illness from COVID-19 including obesity, heart disease, and diabetes.^14,15^ Rural populations also have higher rates of tobacco use, which puts them at greater risk of poor outcomes from this respiratory virus.^15^ Disparities in access to healthcare among residents as well as resource-limited healthcare systems in rural communities also increase the likelihood of higher morbidity and mortality from COVID-19.^10,14^ These factors make it particularly important that college students living in rural areas understand effective ways to reduce their risk of COVID-19 infection and thus prevent spreading of the virus if they were to return home from their college campus.

### Differences in Race and Ethnicity

As cases of COVID-19 continue to rise, alarming data has shown that racial and ethnic minorities have been disproportionately impacted.^16^ This study indicates racial differences in prevention beliefs and practices in college students but results are likely multifactorial. Longstanding institutional and structural inequities, which have played a key role in the racial disparities seen in COVID-19 outcomes, may also play a vital role in racial differences in COVID-19 prevention beliefs and practices in college students. Black or African American, Hispanic/Latino, and Asian students consistently showed greater prevention beliefs and usage of prevention practices in comparison to white or American Indian or Alaska Native/Native Hawaiian or Other Pacific Islander (AIAN/NHOPI) college students. Increased awareness of racial disparities in COVID-19 outcomes may have made prevention practices more of a priority to ethnic and racial minority students. This may explain the heightened fear of personally catching COVID-19 or family members catching the disease that was seen in ethnic and racial minority students in comparison to white students, who showed less concern. Communities of color are also more likely to reside in areas with higher housing density and work essential professions that do not give them the privilege to work from home.^17^ This may make following social distancing regulations extremely difficult, which may explain the increased perceived importance of mask usage and social distancing that was seen in ethnic and racial minority students.

### Limitations

Selection bias and response bias are potential limitations of survey studies and may exist in these results. To minimize selection bias, a study description and link to the survey was emailed to all enrolled undergraduate students at the participating university. With these procedures, our response rate was 22.9% of 21,128 undergraduate students. This is a similar to a response rate cited in a previous study among college students.^18^

Like most survey studies, our results may also be influenced by social desirability bias. Social desirability bias refers to the tendency of respondents to answer in a way that they believe portrays them favorably. College students who are aware that COVID-19 preventative practices are viewed as important by most governing and authoritative bodies may feel that they should respond in ways that are congruent with those views. Because anonymity of responses was stressed to participants, this effect may be less influential on our results.

Students responded to the survey in the summer and before the start of the new academic year. Therefore, many may have been residing in their hometown. As mentioned previously, COVID-19 preventative practices may be influenced by students’ own city or state policies regarding COVID-19 prevention and mitigation. For example, students living in areas with city or state mask mandates would be more likely to respond that they always wear a mask in public, simply because it is required by law. Because of this, we also asked participants their opinions on the level of importance of various COVID-19 prevention strategies. These responses may be less affected by local policies and correspond more with their own beliefs.

## CONCLUSION

Significant differences exist in COVID-19 prevention beliefs and practices in college students based on sex, size of town in which one lives, race, and ethnicity. Interventions designed to educate college students on effective ways of reducing their risk of COVID-19 infection and plans for wide-spread COVID-19 vaccinations should take into consideration these differences.

Prevention of the spread of COVID-19 from the college setting to rural parts of the country is critical when students return home on weekends and holidays. Students from these areas report some of the lowest adherence to prevention guidelines and additional interventions should be developed to increase preventative practices in these students.

A common pattern seen among all demographic groups was a greater concern for family members catching COVID-19 than contracting the virus themselves. It has been well-established that older individuals, particularly those age 65 or older, have higher rates of morbidity and mortality from COVID-19 compared to younger individuals.^19^ Many college students are likely aware of this and may perceive less personal risk of severe disease and death from COVID-19 but worry that older family members may not recover as easily. This concern for morbidity and mortality among older family members may be used an important teaching point when discussing COVID-19 prevention among college students. By emphasizing the importance of preventing viral transmission to family members, college students may be encouraged to reduce their own risk of COVID-19 infection and receive a COVID-19 vaccine when available.

## Data Availability

All data was obtained and is stored on cloud based servers through Qualtrics

## Supplemental Materials

**Supplemental Table.**
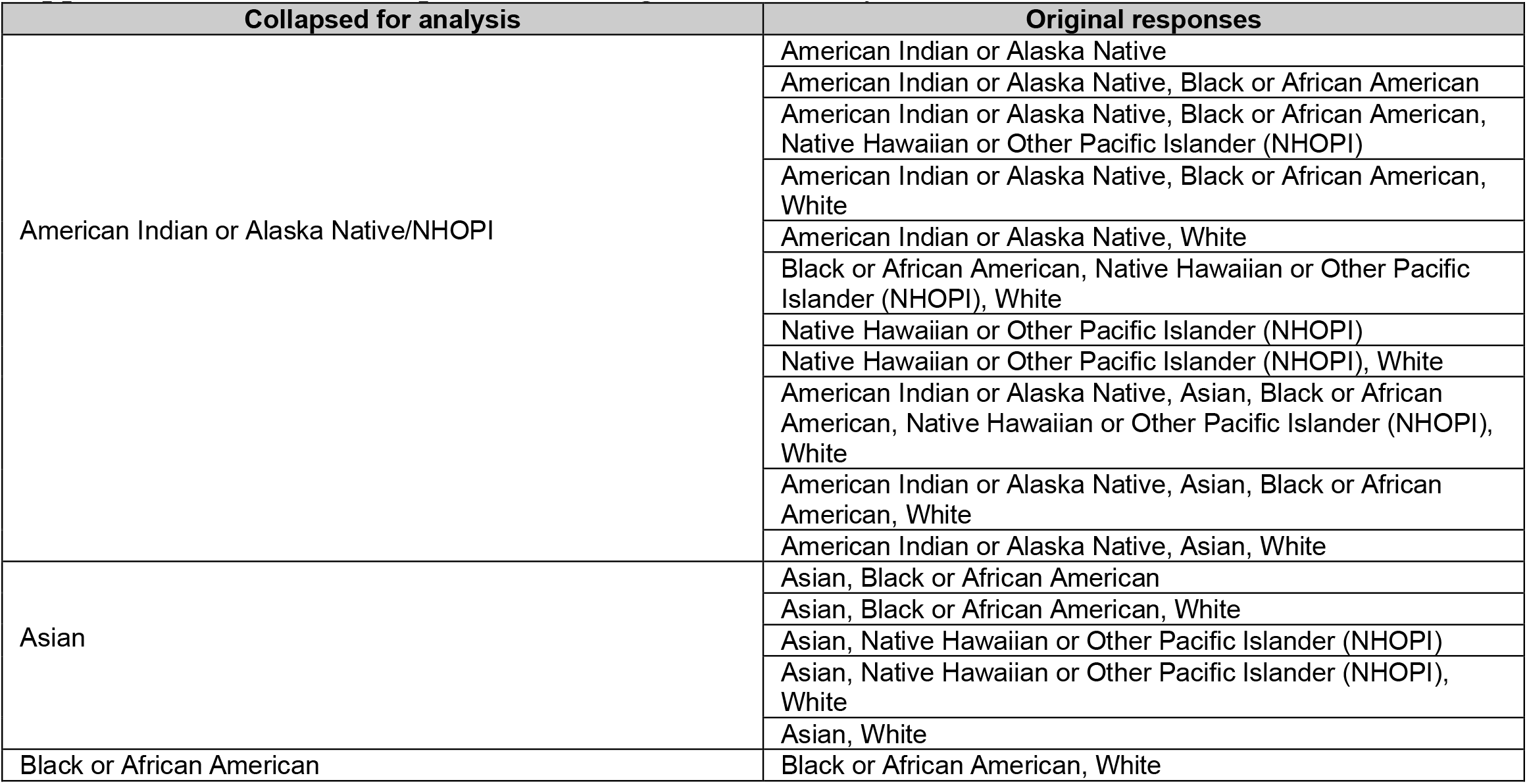
Collapsed race categories for analysis.

| Collapsed for analysis | Original responses |
| --- | --- |
| American Indian or Alaska Native/NHOPI | American Indian or Alaska Native |
|  | American Indian or Alaska Native, Black or African American |
|  | American Indian or Alaska Native, Black or African American, Native Hawaiian or Other Pacific Islander (NHOPI) |
|  | American Indian or Alaska Native, Black or African American, White |
|  | American Indian or Alaska Native, White |
|  | Black or African American, Native Hawaiian or Other Pacific Islander (NHOPI), White |
|  | Native Hawaiian or Other Pacific Islander (NHOPI) |
|  | Native Hawaiian or Other Pacific Islander (NHOPI), White |
|  | American Indian or Alaska Native, Asian, Black or African American, Native Hawaiian or Other Pacific Islander (NHOPI), White |
|  | American Indian or Alaska Native, Asian, Black or African American, White |
|  | American Indian or Alaska Native, Asian, White |
| Asian | Asian, Black or African American |
|  | Asian, Black or African American, White |
|  | Asian, Native Hawaiian or Other Pacific Islander (NHOPI) |
|  | Asian, Native Hawaiian or Other Pacific Islander (NHOPI), White |
|  | Asian, White |
| Black or African American | Black or African American, White |

Survey questions with question logic not shown.

## COVID-19 Prevention Beliefs and Practices in NCAA Athletes and Non-Athlete Students

Q1: What school do you attend?

Q2: Are you of Hispanic, Latino, or Spanish origin?

- No, not of Hispanic, Latino, or Spanish origin
- Yes, Hispanic, Latino, or Spanish

Q3: What is your race? Select all that apply.

- American Indian or Alaska Native
- Asian
- Black or African American
- Native Hawaiian or Other Pacific Islander (NHOPI)
- White

Q4: Since mid-March 2020, in what state have you lived the most?

Q5: What is the population of the town/city where you have lived the most since mid-March 2020?

- 50,000 or more people
- At least 2,500 and less than 50,000
- Less than 2,500

Q6: Are you currently living in the same town as your college/university?

- Yes
- No

Q7: Are you a NCAA student athlete?

- Yes
- No

Q8: Do you compete in Women’s or Men’s NCAA sports?

- Women’s
- Men’s

Q9: What college sport do you play? Select all that apply.

- Baseball
- Basketball
- Beach Volleyball
- Bowling
- Cross country
- Fencing
- Field Hockey
- Football
- Golf
- Gymnastics
- Ice Hockey
- Lacrosse
- Rifle
- Rowing
- Skiing
- Soccer
- Softball
- Swim and Dive
- Tennis
- Track and Field (indoor)
- Track and Field (outdoor)
- Volleyball
- Water Polo
- Wrestling

Q10: What sex were you assigned at birth, meaning on your original birth certificate?

- Male
- Female
- Q11: Which best describes your current gender identity?
- Male
- Female
- Indigenous or other cultural gender minority identity (e.g. two-spirit)
- Something else (e.g. gender fluid, non-binary)

## The following questions are regarding your beliefs and practices on COVID-19 prevention

Q12: Since March 2020, how often have you worn a mask or face covering when going to the grocery store?

- always
- often
- sometimes
- rarely
- never
- I have not been inside a grocery store since March

Q13: Do you think it is important to wear a mask or face covering when going into public places?

- very important
- important
- moderately important
- slightly important
- unimportant

Q14: Are you worried about personally catching COVID-19?

- very worried
- worried
- moderately worried
- slightly worried
- not worried

Q15: Are you worried about a family member catching COVID-19?

- very worried
- worried
- moderately worried
- slightly worried
- not worried

Q16: Over the past 3 months, have you become more or less worried about catching COVID-19?

- much more worried
- slightly more worried
- unchanged
- slightly less worried
- much less worried

Q17: Do you think that it is important to stay 6 feet apart (i.e. social distancing) from people who are not family in the same household or roommates?

- very important
- important
- moderately important
- slightly important
- unimportant

Q18: How often do you try to stay 6 feet apart (i.e. social distancing) from people who are not family or roommates when out in public?

- always
- often
- sometimes
- rarely
- never

Q19: Do you believe it is important to frequently wash your hands with soap and water for at least 20 seconds or use hand sanitizer?

- very important
- important
- moderately important
- slightly important
- unimportant

Q20: Have you received any education or resources on COVID-19 prevention from your athletic department (if a student athlete) or university?

- yes
- no

